# Exome-wide analysis of copy number variation shows association of the human leukocyte antigen region with asthma in UK Biobank

**DOI:** 10.1101/2021.12.15.21267845

**Authors:** Katherine A. Fawcett, German Demidov, Nick Shrine, Megan L Paynton, Stephan Ossowski, Ian Sayers, Louise V. Wain, Edward J. Hollox

## Abstract

**Background:** The role of copy number variants (CNVs) in susceptibility to asthma is not well understood. This is, in part, due to the difficulty of accurately measuring CNVs in large enough sample sizes to detect associations. The recent availability of whole-exome sequencing (WES) in large biobank studies provides an unprecedented opportunity to study the role of CNVs in asthma.

**Methods:** We called common CNVs in 49,953 individuals in the first release of UK Biobank WES using ClinCNV software. CNVs were tested for association with asthma in a stage 1 analysis comprising 7,098 asthma cases and 36,578 controls from the first release of sequencing data. Nominally-associated CNVs were then meta-analysed in stage 2 with an additional 17,280 asthma cases and 115,562 controls from the second release of UK Biobank exome sequencing, followed by validation and fine-mapping.

**Results:** Five of 189 CNVs were associated with asthma in stage 2, including a deletion overlapping the *HLA-DQA1* and *HLA-DQB1* genes, a duplication of *CHROMR/PRKRA*, deletions within *MUC22* and *TAP2*, and a duplication in *FBRSL1*. The *HLA-DQA1, HLA-DQB1, MUC22* and *TAP2* genes all reside within the human leukocyte antigen (HLA) region on chromosome 6. *In silico* analyses demonstrated that the deletion overlapping *HLA-DQA1* and *HLA-DQB1* is likely to be an artefact arising from under-mapping of reads from non-reference HLA haplotypes, and that the *CHROMR/PRKRA* and *FBRSL1* duplications represent presence/absence of pseudogenes within the HLA region. Bayesian fine-mapping of the HLA region suggested that there are two independent asthma association signals. The variants with the largest posterior inclusion probability in the two credible sets were an amino acid change in *HLA-DQB1* (glutamine to histidine at residue 253) and a multi-allelic amino acid change in *HLA-DRB1* (presence/absence of serine, glycine or leucine at residue 11).

**Conclusions:** At least two independent loci characterised by amino acid changes in the *HLA-DQA1, HLA-DQB1* and *HLA-DRB1* genes are likely to account for association of SNPs and CNVs in this region with asthma. The high divergence of haplotypes in the HLA can give rise to spurious CNVs, providing an important, cautionary tale for future large-scale analyses of sequencing data.

## Background

Asthma is a chronic, inflammatory lung condition affecting over 300 million people worldwide. The proportion of population variance in asthma risk attributable to genetic variation has been estimated to be between 35 and 95% (1), and about 200 genetic loci have been associated with asthma (2). However, the variants discovered to date only account for a small proportion of heritability (2), and unmeasured genomic structural variants such as copy number variants (CNVs) may also contribute to genetic risk. Indeed, CNVs have been shown to play a role in a number of common, complex diseases and traits (3). There is strong evidence that CNVs are also important contributors to asthma risk (4-6), but to date there have been few reported associations with specific structural variants, including CNVs.

Previous studies of genome-wide CNVs in common, complex disease have detected CNVs using hybridisation-based techniques such as SNP genotyping arrays. However, these methods have limited genome coverage due to variability in SNP density, and only have the resolution to detect the largest CNVs reliably (7). The increasing availability of large high-throughput sequencing datasets offers an unprecedented opportunity to investigate a more comprehensive set of CNVs and other structural variants. The UK Biobank, a population-based cohort of half a million volunteer participants, released exome sequencing data on approximately 50,000 participants deliberately enriched for individuals with asthma in March 2019 (8). They released a second tranche of exome sequencing, including an additional approximately 150,000 individuals, at the end of 2020 (9). This resource allows researchers to detect exome-wide CNVs and test then for association with asthma, potentially identifying novel genetic drivers of asthma and new mechanistic insights at asthma-associated loci.

In this study, we detected CNVs affecting exons in 7,098 asthma cases and 36,578 controls from UK Biobank and tested them for association with asthma status. We then performed meta-analyses of asthma-associated CNVs from this first stage with and an additional set of 17,280 asthma cases and 115,562 controls from the second tranche of UK Biobank exome sequencing. *In silico* validation of CNVs demonstrating reproducible association with asthma was sought in publicly available datasets (including those with long-read sequencing data), and the causal role of validated CNVs in asthma was investigated.

## Methods

### Study participants

The UK Biobank study is described here: https://www.ukbiobank.ac.uk/. Participants were included in this study if they were in the first or second tranche of exome sequencing data (8, 9), were of genetically inferred European ancestry, and were not first- or second-degree relatives of anyone already selected for inclusion.

Individuals were defined as having asthma if they either self-reported doctor-diagnosed asthma (fields 6152 or 22127) or had an International Classification of Diseases (ICD)10 code for asthma (J45* or J46*) in hospital inpatient records. Individuals were defined as controls if they had no self-reported doctor-diagnosed asthma, no ICD10 code for asthma in hospital inpatient records, and did not report asthma in an interview with a nurse (field 20002). Cases and controls reporting chronic bronchitis or emphysema (fields 6152, 22128, or 22129) or with an ICD10 code for chronic bronchitis or emphysema in hospital inpatient records were excluded from the analysis.

The numbers of individuals remaining in the first and second tranche of exome sequencing data at each stage of selection are given in Figure 1.

**Figure 1.**
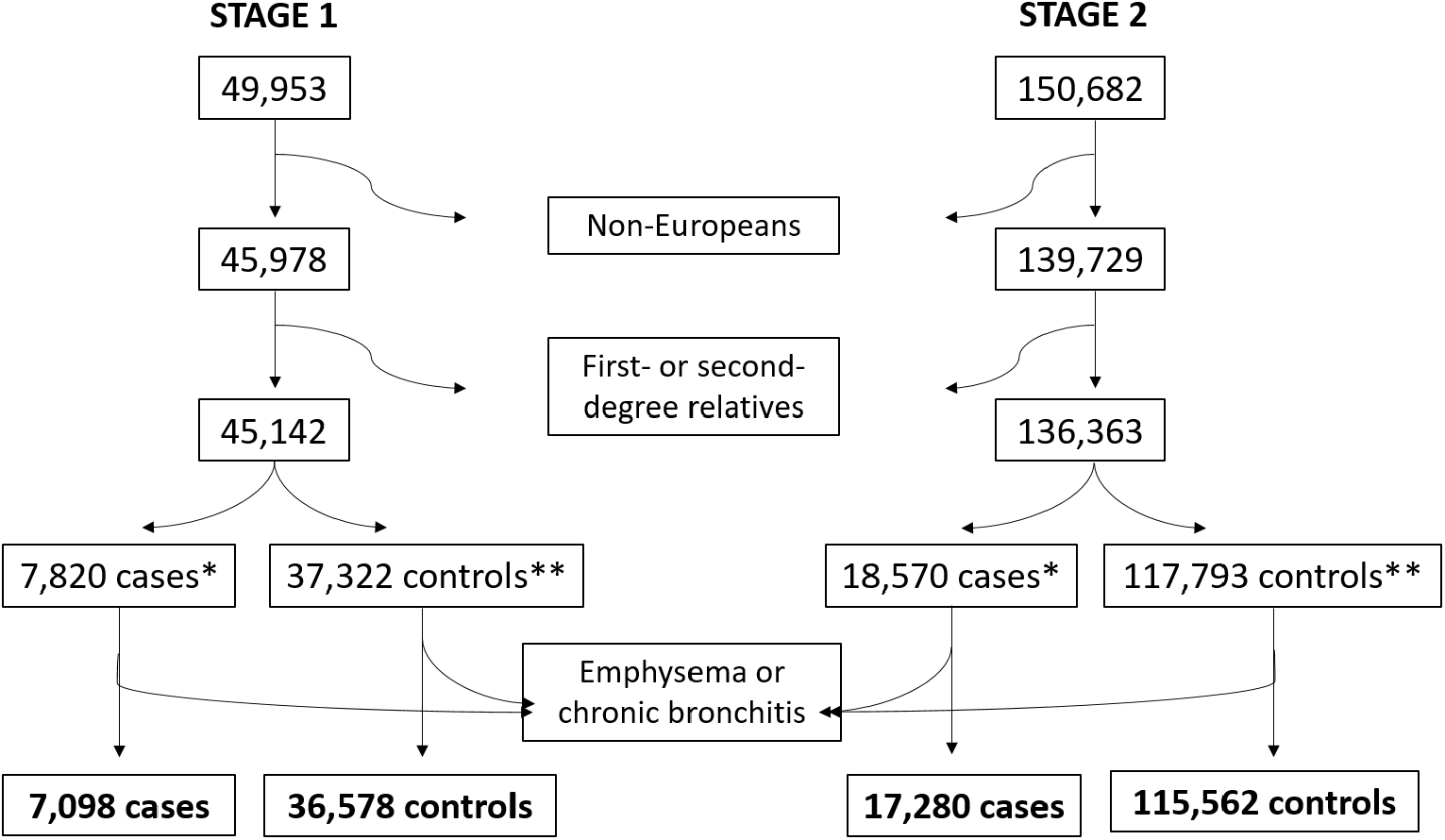
Number of UK Biobank participants meeting study selection criteria. *Asthma cases defined as having self-reported doctor-diagnosed asthma or an ICD10 code for asthma in hospital inpatient records. **Controls defined as having no self-reported doctor-diagnosed asthma, no ICD10 code for asthma, and no self-reported asthma from nurse’s interview.

### Exome sequencing

Exome sequencing data was available from UK Biobank for 200,635 individuals (49,953 from the first release and an additional 150,682 from the second release). Exomes were captured using the IDT xGen Exome Research Panel v1.0 including supplemental probes prior to 75bp paired end sequencing on the Illumina NovaSeq 6000 platform using S2 (first tranche) and S4 (additional samples in the second tranche) flow cells. For the first tranche of exome sequencing (our stage 1 set) we used data from the March 2020 release of the UK Biobank exome data (8). These data had been processed using the SPB pipeline (8). For the second tranche of exome sequencing (our stage 2 set) we used data from the December 2020 release processed using the OQFE protocol, which maps sequencing reads to the full GRCh38 reference version including all alternative contigs in an alt-aware manner (9).

### Copy number variant calling and genotyping

Prior to CNV calling, we calculated read depth over exome capture regions from UK Biobank CRAM files using ngs-bits (https://github.com/imgag/ngs-bits) BedCoverage function and a minimum mapping quality of 5. We then used ClinCNV (https://github.com/imgag/ClinCNV) for the detection of medium to large (>=1 exome capture region) germline CNVs, including deletions, duplications and multi-allelic CNVs, based on the principle of depth-of-coverage as described in Additional file 1.

For each call, ClinCNV generated plots of normalised coverage colour-coded by copy number assignment. We performed manual inspection of each call to select those that exhibited distinct copy number clusters. CNVs were annotated using AnnotSV.

### Benchmarking

It can be challenging to assess the accuracy of CNV callers due to the absence of truth sets within the tested cohort. However, we had previously directly measured copy number at the *CCL3L1* locus using paralogue ratio tests in a subset of approximately 5000 UK Biobank participants of European ancestry (10), with copy numbers ranging from 0 to 5. We compared the distribution of copy numbers in these ∼5000 participants to the distribution of copy numbers inferred by ClinCNV in the 49,953 participants from the first release of exome sequencing. For 412 individuals that were genotyped in both studies, we also compared the genotypes from experimental studies with genotypes assigned by ClinCNV for the *CCL3L1* locus.

To assess false negative rates, we downloaded the structural variant calls from phase 3 of the 1000 genomes project (http://ftp.1000genomes.ebi.ac.uk/vol1/ftp/phase3/integrated_sv_map/supporting/GRCh38_positions/ALL.wgs.mergedSV.v8.20130502.svs.genotypes.GRCh38.vcf.gz). We filtered these 1000 genomes variants for copy number changes (SVTYPE = DEL, DUP, CNV, DEL_ALU, DEL_LINE1, or DEL_SVA) with allele frequency in Europeans greater than 5%, and which overlapped one or more UK Biobank exome capture regions by at least 70%. We considered these 1000 genomes CNVs to have been called by ClinCNV in our UK Biobank cohort if the two calls shared alternative alleles with a frequency within 5% of each other.

### Statistical analysis

We took a two-stage approach to identifying CNVs associated with asthma. We first performed an association analysis in individuals from the first tranche of exome sequencing (including 7,098 asthma cases and 36,578 controls). This stage 1 set produced a long-list of CNVs nominally associated with asthma (P<0.05). We then meta-analysed this long-list of CNVs with additional samples from the second tranche of exome sequencing (stage 2) (including 17,280 asthma cases and 115,562 controls). In the stage 2 meta-analysis we used a Bonferroni-corrected overall significance threshold of P<2.65×10^−4^.

To test each copy number variant for association with asthma, logistic regression models were implemented in R, with copy number, sex, age at recruitment, squared age at recruitment, and the first twenty principal components of genome-wide SNP genotypes, measured using the UK Biobank Axiom Genotyping array (to correct for population structure) included as covariates. Conditional analyses were performed by including the variants of interest in the regression model as covariates. LocusZoom was used to generate region plots.

Sensitivity analyses were performed to assess whether the effect of CNVs on asthma risk was altered when allergies were excluded. Individuals with self-reported doctor-diagnosed hayfever, allergic rhinitis and eczema (field 6152) were excluded from cases and controls and the association analysis described above was repeated.

### Power calculations

In stage 1, we had over 80% power to detect an odds ratio of 1.06 with a CNV with an alternate allele frequency of 0.3. Even for low frequency CNVs (alternate allele frequency of 0.05) we had over 80% power to detect an odds ratio of 1.13. Power calculations were carried out using Quanto.

### *In silico* validation of CNVs

CNVs that showed association with asthma were followed up by visualising the read data in the Integrative Genomics Viewer (IGV) (Broad) and checking for related pseudogenes that might lead to spurious CNV calls in resources such as the NCBI gene database (https://www.ncbi.nlm.nih.gov/gene/). We also checked for the presence of the CNVs in studies that used alternative bioinformatics or experimental approaches to identify structural variants (11-13), and using public online repositories such as the Broad CNV Browser (http://www.broadinstitute.org/software/genomestrip/mcnv_supplementary_data) (14), the Database of Genomic Variation (http://dgv.tcag.ca/dgv/app/home) and the genome aggregation database (https://gnomad.broadinstitute.org/). Specifically, if the CNVs we detected in UK Biobank overlapped a CNV of the same type that had a similar frequency in publicly available data then this was considered to be evidence that the CNV was real.

### HLA region fine-mapping

For UK Biobank participants that had SNP data passing quality control (N = 487,409), we re-imputed classical HLA genotypes and constituent predicted amino acid changes within HLA genes. For imputation, we used IMPUTE2 v2.3.2, the UK Biobank haplotypes (Category 100319) as the input and the T1DGC reference panel (containing haplotypes for 5,225 samples from the Type 1 Diabetes Genetics Consortium (T1DGC)) (15). These imputed genotypes were then used alongside imputed SNPs and CNVs of interest to fine-map the signals of genetic association within the HLA region using a Bayesian method (Susie) (16). We restricted fine-mapping to variants with a minor allele frequency greater than 1% within the UK Biobank cohort. Summary statistics from the logistic regression were passed to the susie_rss function with a variant correlation matrix generated using Plink v1.9. Default parameters were used for the Susie analysis. Association of HLA region variants was plotted using Locuszoom. While imputed amino acid changes were included in the fine-mapping, they were excluded from the plots (unless they were present in a credible set) as the presence/absence alleles for all variants in a codon are ascribed the same chromosomal position, which disrupts Locuszoom plotting.

## Results

In the first tranche of UK Biobank exome sequencing (N = 49,953), we used ClinCNV software to call a total of 665 CNVs. Of these CNV calls, 189 showed distinct and well-separated clusters upon visual inspection. Benchmarking showed that ClinCNV calls a high proportion (62.5%) of the common CNVs present in phase 3 of the 1000 genomes project, and that it is capable of accurately inferring copy number at complex multi-allelic loci (Additional file 1: Supplementary Results and Figure S1, Additional file 2: Table S1 and S2).

### Testing CNVs for association with asthma

After exclusion of non-European individuals and relatives, data were available for 7,098 cases and 36,578 controls in the stage 1 cohort, and for 17,280 cases and 115,562 controls in the independent stage 2 cohort (Figure 1). Baseline characteristics for these cohorts are shown in Table 1.

**Table 1.** Baseline characteristics of stage 1 and stage 2 UK Biobank cohorts.

| Trait | Stage 1 |  | Stage 2 |  |
| --- | --- | --- | --- | --- |
|  | Cases (N = 7,098) | Controls (N = 36,578) | Cases (N = 17,280) | Controls (N = 115,562) |
| **Age at recruitment, years** | 56 (8.09) | 56.9 (7.89) | 55.9 (8.23) | 56.7 (8.02) |
| **Female** | 4199 (59.2) | 19561 (53.5) | 10197 (59.0) | 63424 (54.9) |
| **Male** | 2899 (40.8) | 17017 (46.5) | 7083 (41.0) | 52138 (45.1) |
| **FEV<sub>1</sub>, % predicted** | 85.7 (16.6) | 94.0 (14.3) | 88.2 (15.3) | 94.2 (14.1) |
| **FEV<sub>1</sub>/FVC** | 0.733 (0.0778) | 0.769 (0.0551) | 0.741 (0.0709) | 0.769 (0.0555) |
| **Ever smoker** | 3093 (43.6) | 16055 (43.9) | 7590 (43.9) | 50413 (43.6) |
| **Never smoker** | 3901 (55.0) | 20002 (54.7) | 9357 (54.1) | 63253 (54.7) |
| **Unknown** | 104 (1.5) | 521 (1.4) | 333 (1.9) | 1896 (1.6) |
| **Hayfever, rhinitis or eczema status:** |  |  |  |  |
| **Yes** | 3483 (49.1) | 8228 (22.5) | 7727 (44.7) | 23269 (20.1) |
| **No** | 3615 (50.9) | 28350 (77.5) | 9553 (55.3) | 92293 (79.9) |

Data are mean (SD) or N (%), FEV<sub>1</sub> = forced expiratory volume in 1 second, FVC = forced vital capacity

We tested 189 high-quality CNVs for association with asthma in the stage 1. Seventeen CNVs showed nominal association with asthma (P<0.05) (Table 2, Additional file 2: Tables S3 and S4) and were taken forward to stage 2.

**Table 2.**
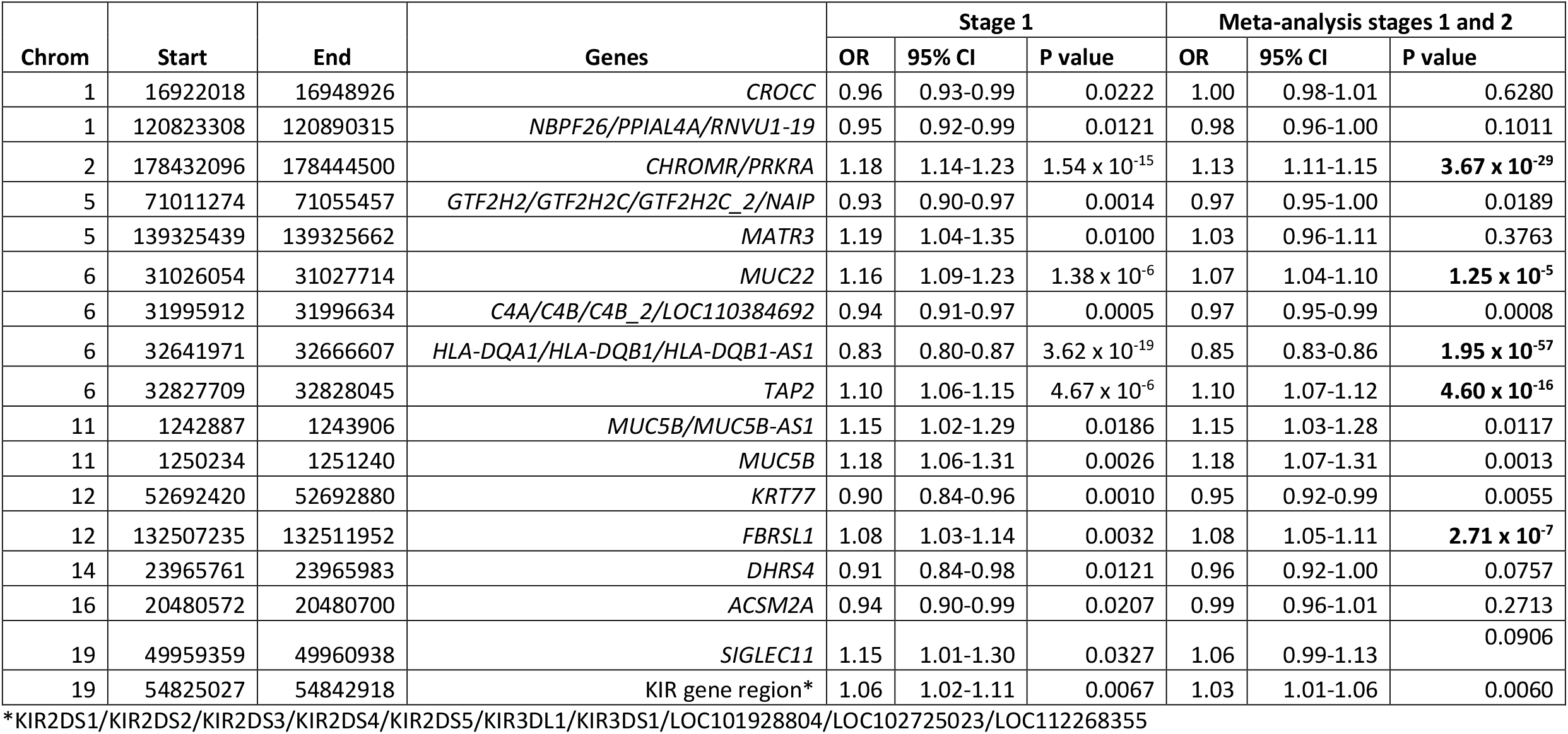
Association of copy number variants with risk of asthma in UK Biobank.

In a meta-analysis of stage 1 samples and the additional independent 17,752 cases and 115,562 controls from the stage 2 cohort, we detected five CNVs associated with asthma at a Bonferroni-corrected P value threshold (P<2.65×10^−4^) (Table 2). Cluster plots are shown in Additional file 1: Figure S2. The CNV with the strongest association signal is predicted to encompass exons 3-5 of the *HLA-DQA1* gene and all of the *HLA-DQB1* gene. These genes are adjacent to each other within the human leukocyte antigen (HLA) region on chromosome 6p21, and SNPs in these genes have been shown to be strongly associated with asthma (17-20).

Of the remaining replicated CNVs, two affect genes also residing in the HLA: a partial deletion of the large, central exon of *MUC22*, and a small deletion within the 3’UTR of *TAP2*. Genetic variants in the *MUC22* gene region (21, 22) and in *TAP2* (2) have been previously associated with asthma and asthma-related traits. The final two asthma-associated CNVs are partial gene duplications on chromosome 2 (affecting exons 4-8 of the *PRKRA* gene and the 3’ end of the *CHROMR* long non-coding RNA gene) and chromosome 12 (affecting exon 2 of the *FBRSL1* gene). These genes have not, as far as we are aware, been genetically associated with asthma before.

All five asthma-associated CNVs showed consistent effect sizes in a sensitivity analysis that excluded cases and controls with common allergic conditions: *HLA-DQA1/HLA-DQB1* (0.87 [0.84-0.89], p = 1.20 × 10^−24^), *PRKRA* (1.13 [1.10-1.16], p = 4.25 × 10^−17^), *MUC22* (1.04 [1.00-1.08], p = 0.048), *TAP2* (1.09 [1.06-1.12], p = 3.12 × 10^−8^), and *FBRSL1* (1.08 [1.04-1.12], p = 0.0001).

### Validation and characterisation of asthma-associated CNVs

To seek validation for our five CNVs associated with asthma and to identify putative breakpoints, we examined the mapped reads in the UK Biobank CRAM files using IGV and searched for our asthma-associated CNVs in publicly available short- and long-read sequencing results.

#### MUC22 and TAP2

Examination of the mapped reads in the UK Biobank CRAM files within the *MUC22* and *TAP22* CNV regions showed no reads in individuals genotyped as homozygous for the deletion, as expected (Additional file 1: Figure S3). The *MUC22* and *TAP2* deletions were identified in long-read data reported by Audano et al. (11) (hg38 coordinates chr6: 31026230-31027303 and chr6: 32827728-32827904 respectively) and in Chaisson et al. (12) (hg38 coordinates chr6: 31026229-31027304 and chr6: 32827726-32827903 respectively), and in the 1000 genomes dataset (hg38 coordinates: 31026238-31027306 and chr6:32827726-32827903 respectively). The allele frequency of the deletion in individuals of European ancestry from 1000 genomes (0.145 and 0.269 for *MUC22* and *TAP2* respectively) approximately matches the frequency amongst the UK Biobank participants (0.115 and 0.244 respectively, Additional file 1: Table S4). It is therefore likely that these CNV calls represent real deletions within the *MUC22* and *TAP2* genes.

#### HLA-DQA1/HLA-DQB1

The whole-exome sequencing data from individuals called as homozygous for the *HLA-DQA1/HLA-DQB1* deletion contained mapped reads within the reported CNV boundaries (Additional file 1: Figure S4). Mapped reads were also present within the reported CNV boundaries in pilot whole-genome sequencing (https://biobank.ndph.ox.ac.uk/showcase/field.cgi?id=23183) from the same individuals (Additional file 1: Figure S4). These reads exhibited greater sequence divergence from the primary reference sequence compared to the reads in individuals without the deletion, and divergent reads showed supplementary alignments to alternative chromosome 6 and HLA sequences within the full hg38 reference. HLA haplotypes show extremely high sequence divergence across the *DQA* and *DQB* genes (23). This high sequence divergence both from other haplotypes and the reference sequence potentially limits mappability of sequencing reads from divergent haplotypes onto the reference genome, leading to underestimation of the number of reads derived from that region. The apparent deletion at this locus was therefore likely to be due to under-mapping of reads from reference-divergent HLA haplotypes.

To investigate this further, we imputed HLA alleles in UK Biobank and found that the *HLA-DQA1/DQB1* CNV was almost perfectly correlated with the *HLA-DQA1*\*01 type (Spearman correlation = 0.97). Those without the deletion were homozygous for the *HLA-DQA1*\*01 type and those heterozygous or homozygous for the deletion were heterozygous or homozygous for non-*HLA-DQA1*\*01 types respectively.

This CNV was not found in publicly available long-read sequencing results. However, there is evidence for a CNV overlapping the *HLA-DQA1* and *HLA-DQB1* genes on the Broad CNV browser for the Handsaker et al. study (14) (CNV_M1_HG19_6_32603984_32627361) but, as this call was based on read depth as well, it is likely to suffer from the same artefacts.

The artefactual nature of this CNV may account for the large departure of the genotype data from Hardy Weinberg Equilibrium (Additional file 2: Table S4).

#### CHROMR/PRKRA and FBRSL1

Individuals with duplications in the *PRKRA* gene showed read pairs spanning exon-exon boundaries, whereas those without the duplication did not (Additional file 1: Figure S5). These exon-spanning reads also have secondary alignments to the alternative chromosome 6 and HLA sequences. Previous work has shown that the HLA region DR53 haplotype contains an intronless, retrotransposed *PRKRA* pseudogene (also referred to as *PRKRAP1*) proximal to the *HLA-DRB7* pseudogene on GL000253v2_alt and GL000256v2_alt GRCh38 sequences (24). This suggests that the intron-spanning read-pairs mapping to *PRKRA* might actually arise from the pseudogene, resulting in an apparent increase in copy number of *PRKRA* in individuals carrying HLA haplotypes containing the pseudogene. The association with asthma is therefore not necessarily with the *PRKRA* gene but potentially with HLA variation in linkage disequilibrium (LD) with the presence/absence of the pseudogene. The background noise of reads arising from the canonical *PRKRA* gene, on top of the reads arising from the pseudogene, probably accounts for why ClinCNV struggles to classify individuals into copy number bands at this locus (Additional file 1: Figure S2A). As can be seen in the cluster plots, some individuals clustering with the copy number of 4 group, are nonetheless assigned a copy number of 3, hence the large departure from Hardy-Weinberg Equilibrium (Additional file 2: Table S4).

Similarly, individuals with the *FBRSL1* gene duplication exhibit read pairs spanning exon boundaries (Additional file 1: Figure S5) and these reads map to alternative HLA sequences. A recent study identified a *FBRSL1* processed pseudogene on chromosome 6 (25), suggesting that this CNV also represents variation in the HLA region.

Further evidence that the *PRKRA* and *FBRSL1* signals are due to causal variation in the HLA region is provided by linkage disequilibrium and conditional analyses with HLA variants. Both CNVs were correlated with variation in the HLA region, but not their supposedly surrounding SNPs on chromosomes 2 and 12 respectively. The associations of *PRKRA* and *FBRSL1* CNVs with asthma were also abolished by conditioning on certain HLA variants (Additional file 2: Table S5). For example, the *PRKRA* CNV association was abolished by conditioning on various amino acid changes in the *HLA-DRB1, HLA-DQA1* and *HLA-DQB1* genes, and the *HLA-DQA1*\*01 allelotype. Likewise, the *FBRSL1* CNV association was abolished by including *HLA-C*\*07:01, *HLA-B*\*08 and *HLA-B*\*08:01 types in the model, as well as amino acid changes in *HLA-C, HLA-B, HLA-DRB1, HLA-DQA1*, and *HLA-DQB1*.

### Fine-mapping the HLA region

Having established that all our reproducible signals derive from the HLA region, we performed fine-mapping of this region using all imputed SNPs/indels (N = 19,891), imputed HLA alleles (N = 136) and amino acid changes (N = 835) with minor allele frequency greater than 1%, as well as the *MUC22* CNV, the *TAP2* CNV and the presence of *PRKRA* and *FBRSL1* pseudogenes, in UK Biobank. Using a Bayesian fine-mapping method (Susie), we identified two credible sets over the *HLA-DRB1, HLA-DQA1*, and *HLA-DQB1* genes (Figure 2, credible set 1 variants shown in red circles and credible set 2 variants shown in blue circles). The first set contained 66 variants and the variant with the largest posterior inclusion probability was rs1140343 (Additional file 2: Table S6), a missense change leading to substitution of a histidine for glutamine at residue 253 of *HLA-DQB1*. However, due to the large number of variants in this credible set, the top variant had a modest posterior inclusion probability of 0.082. Presence/absence of arginine at residue 55 was also in the top 5 variants with a posterior inclusion probability of 0.044, and this variant actually had the largest effect size and most significant p value (Additional file 2: Table S6). The second set contained 6 variants and the top variant, with a posterior inclusion probability of 0.555, was presence/absence of the amino acids serine, glycine or leucine at residue 11 in *HLA-DRB1*.

**Figure 2.**
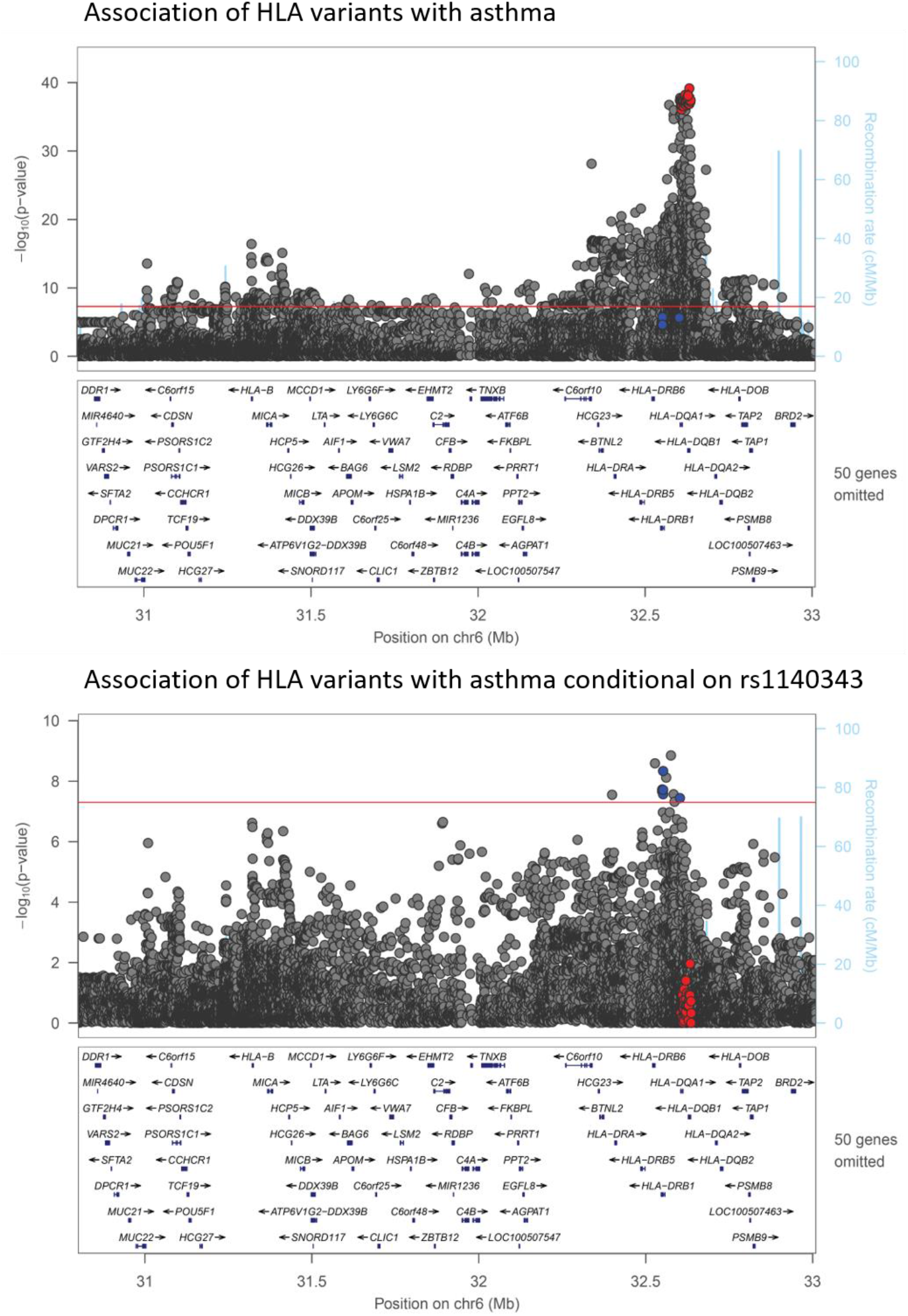
Association of HLA variants with asthma in UK Biobank. Locuszoom plots of the HLA region showing association of HLA region variants with asthma (top panel) and association of HLA region variants with asthma conditional on the SNP with the top posterior inclusion probability in credible set 1 (bottom panel). Variants in credible set 1 are highlighted in red and variants in credible set 2 are highlighted in blue.

The variants in credible set 1 were amongst the most significantly associated with asthma (red circles in top panel of Figure 2), while the variants in credible set 2 do not reach genome-wide significance (P<5×10^−8^, blue circles in top panel of Figure 2). However, when rs1140343 (the SNP with the top PIP in credible set 1) is added to the regression model, the variants in credible set 2 are amongst the most associated with asthma (bottom panel of Figure 2). There are two variants not present in credible set 2 that exhibit greater statistical association with asthma after adjustment for rs1140343. These variants show only modest correlation with the top variants from credible set 2 (r^2^ = 0.496 and r^2^ = 0.209 respectively).

No CNV or pseudogene signals were part of these credible sets, suggesting that these are not the underlying causal variants.

## Discussion

We have called CNVs using exome sequencing data in over 200,000 individuals from the UK Biobank study. Out of 189 putative CNV calls showing good separation between copy number clusters, five were reproducibly associated with asthma, including three deletions within the HLA region on chromosome 6, a duplication affecting the *CHROMR*/*PRKRA* genes on chromosome 2 and a duplication affecting the *FBRSL1* gene on chromosome 12. Visual inspection of mapped reads from exome sequencing and examination of publicly-available data showed that the CNV showing the strongest association with asthma, overlapping the *HLA-DQA1* and *HLA-DQB1* genes, was likely to be an artefact of under-mapping of reads from reference-divergent HLA haplotypes, and that the duplications affecting the *CHROMR*/*PRKRA* and *FBRSL1* genes were both likely to be artefacts of the polymorphic presence/absence of processed pseudogenes within the HLA region. Fine-mapping of imputed HLA variation and putative CNVs demonstrated that there are likely to be at least two real, independent, association signals for asthma within the HLA region, one involving primarily *HLA-DQA1* and *HLA-DQB1* variation and one involving primarily *HLA-DRB1* variation. The top variants within the credible sets are missense amino acid changes within the *HLA-DQB1* and *HLA-DRB1* genes respectively. The putative HLA CNVs were not present in either of the credible sets representing these signals, suggesting that they are not responsible for the association of HLA variation with asthma.

The HLA region has long been known to play an important role in asthma pathogenesis, presumably through the role of HLA genes in regulating immune processes. Indeed, the *HLA-DQ* locus was the first genetic locus to be associated with asthma (26). Since then, many genetic studies have identified associations between HLA genes and susceptibility to asthma (2, 18, 20, 27), as well as asthma subtypes (17, 19, 28, 29) and related traits such as serum IgE levels (30, 31). Our fine-mapping analysis suggest that there are at least two independent genetic risk loci for asthma within the HLA region. The first signal, represented by credible set 1, contains variants in and around the *HLA-DQA1* and *HLA-DQB1* genes. The variant with the highest posterior inclusion probability in credible set 1 changes amino acid 253 (predicted to lie within the cytoplasmic domain) in the *DQB1* gene from glutamine to histidine. All the variants in credible set 1 are closely correlated and are also in linkage disequilibrium with previously reported asthma variants. For example, the Q253H amino acid change (rs1140343) is correlated with rs9273349, the *HLA-DQ* signal from the first GWAS of asthma (18) (r2 = 0.813). The variant with the highest posterior inclusion probability in credible set 2 is presence/absence of the amino acids S, G, or L at residue 11. This residue is reported to lie in the P4 peptide binding pocket of *HLA-DRB1* (32) and amino acid changes at this position have previously been associated with autoimmune conditions such as rheumatoid arthritis, type 1 diabetes, and systemic lupus erythematosus (32-34). As far as we are aware, this variant is not well-correlated with previous asthma signals.

Detection of CNVs in large sequencing datasets such as the UK Biobank has only become feasible in the last few years and we will see increasing numbers of publications based on these data. In our study, the presence of pseudogenes in the HLA region led to apparent (artefactual) associations with regions on chromosome 2 and 12. Moreover, it is likely that the top CNV overlapping the *HLA-DQA1* and *HLA-DQB1* genes is an artefact of under-mapping of reads from reference-divergent HLA haplotypes. This demonstrates the pitfalls of using short-read sequencing to call CNVs and the importance of validating CNV calls in independent datasets.

We acknowledge several limitations of this study. First, we had over 80% power to detect modest effect sizes from common CNVs in our stage 1 analysis, but might have missed modest effect sizes from lower frequency CNVs (we had under 80% power to detect odds ratios of less than 1.13 for variants with allele frequency of 0.05). Second, our benchmarking suggested that at least 62.5% of common copy number variants from phase 3 of the 1000 genomes project were identified by our CNV calling algorithm. It is therefore possible that there are undetected CNV association signals for asthma. Third, some of our asthma-associated CNVs from the stage 1 analysis exhibited poor quality genotyping in the stage 2 cohort and therefore lack of association of these CNVs should be interpreted cautiously. Fourth, we have not comprehensively assayed CNVs across the genome or frequency spectrum. Future work will include analysis of whole-genome sequencing data soon to be released by UK Biobank and identification of rare CNVs.

## Conclusions

Our data suggests that common CNVs detectable from exome sequencing, and at least one exon in length, are unlikely to be as important for asthma susceptibility as SNP loci. All the asthma-associated CNVs identified in this study represent variation in the HLA region. We showed how the high divergence of haplotypes in the HLA region can give rise to spurious CNVs, providing an important, cautionary tale for future large-scale analyses of sequencing data. Fine-mapping the HLA region suggested that there are at least two asthma association signals in this region and that amino acid changes in *HLA-DQA1, HLA-DQB1* and *HLA-DRB1* genes are most likely to be the underlying causal variations.

## Data Availability

All data (summary statistics) generated or analysed during this study are included in this published article [and its supplementary information files]. The ClinCNV genotyping will be available to approved UK Biobank researchers as a returned dataset.

## List of abbreviations

CNV: Copy number variant
WES: Whole-exome sequencing
SNP: Single nucleotide polymorphism
IGV: Integrative Genomics Viewer
HLA: Human Leukocyte Antigen
T1DGC: Type 1 Diabetes Genetics Consortium

## Declarations

### Ethics approval and consent to participate

This study used anonymised data from UK Biobank, which comprises over 500,000 volunteer participants aged 40–69 years recruited across Great Britain between 2006 and 2010. The protocol and consent were approved by the UK Biobank’s Research Ethics Committee. Our analysis was conducted under approved UK Biobank data application number 56607.

### Consent for publication

Not applicable.

### Authors’ contributions

KAF was responsible for the design of the study, the acquisition of data under approved UK Biobank application 56607, the analysis and interpretation of the data, and the writing of the manuscript. GD created the ClinCNV software used to call and genotype CNVs, and also contributed to the analysis and interpretation of the data, and the writing of the manuscript. NS and MLP imputed HLA alleles in UK Biobank, used in conditional analyses and the fine-mapping part of this study. SO supervised GD to create the ClinCNV software. IS, LVW, and EJH contributed to the study design and interpretation of the results, and also the revision of the manuscript. All authors read and approved the final manuscript.

## Acknowledgements

We would like to acknowledge the UK Biobank and all the participants for generating this important health research resource. This study used the ALICE and SPECTRE High Performance Computing Facilities at the University of Leicester.

## Additional files

Additional file 1 (.docx) Supplementary Methods, Results and Figures. This file contains additional details of the ClinCNV method, additional details of benchmarking results, and Figures S1-5.

Additional file 2 (.xlsx) Supplementary Tables. This file includes Tables S1-6.

