## Additional file 1 for "Exome-wide analysis of copy number variation shows association of the human leukocyte antigen region with asthma in UK Biobank"

**Supplementary Methods: ClinCNV**

*Data Normalization*

ClinCNV utilizes coverage depth values measured within those regions targeted by exome capture prior to sequencing, corrected for library size, GC content and length of targeted regions using rolling median as an estimator for normalization values. A squared root transformation is applied for variance stabilization. Furthermore, ClinCNV removes regions with coverage patterns that show a very low raw read coverage across the cohort or regions with extreme GC content. Taking into account various technical factors that appear to influence the genome-wide coverage distribution, ClinCNV subdivides the cohort into groups of samples with similar coverage profile. UMAP dimension reduction and further DBSCAN is used with the distance defined as Pearson correlation between coverage profiles smoothed by averaging of 10 adjacent regions. The optimal number of clusters has been selected in order to maximize similarity within groups, while having at least 50 samples within each cluster required for accurate estimation of statistical model parameters. Finally, ClinCNV normalizes the corrected coverage of each tile within each group with its mode found by kernel density estimation for multi-allelic CNVs (defined as more than 5% of samples are not diploid at the site of multi-allelic CNV) calling.

*Statistical model inference*

For each window we constructed one statistical model for each of the expected copy-number states. Since the mode of coverage used on the final normalization step estimates the expected value of the major allele's coverage, the coverage of samples having the major allele genotype will be centred on 1. For each particular CN change we know the expected location shift (i.e. sqrt(0.5) for heterozygous deletions within a normally disomic region). We assume that the variance of a region's coverage is equal for all copy numbers except homozygous deletions, since we used variance-stabilized counts. Thus we estimate the variance for each window using robust estimators and model different copy numbers as normally distributed random variables with different, but *a priori* known expected values. For windows that have a non-diploid major allele we make the additional assumption that minor allele frequency (MAF) is greater than 5%, since location shifts of copy number changes in case of high copy number major alleles become small and indistinguishable from the natural variability. Hence, we can detect such deviations only if they are supported by sufficient number of samples. We used constrained mixture modelling analysis for variants with MAF>5% and additionally assumed that the majority of samples show a copy-number of 0 to 16 at such sites.

*CNV Calling*

Subsequent to statistical model estimation we calculate matrices of likelihoods for all tiles in all samples and all CN models. Thus, we will have a matrix of likelihoods of size (number of distinct copy number states * number of targeted regions). First, we select the most probable state (e.g. the state that models the most common copy number state with the assumption that other copy numbers are present with MAF<2.5%) as the baseline. Then, we search the segment (group of adjacent tiles) with the largest evidence of an alternative state within the borders of the investigated interval. Initially we start with a full chromosome arm as such interval. To this end we subtract the interval tile's log-likelihoods under the baseline model $\log(L(M_b))$ from the log-likelihoods under the alternative state's models $\log(L(M_a))$. The log-likelihood difference becomes negative if the observed data is more likely under the assumptions of the baseline model and positive if the data supports the alternative model. Thus we solve the maximum sub-array problem (1) for each alternative state $M_{a0}$, $M_{a1}$, etc., separately and find the segment with the maximum positive log-likelihood difference for each state.

After choosing the state with the maximum logarithm of likelihood ratio $2\log(LR(M_a,M_b))=2(\log(L(M_a)) - \log(L(M_b)))$ we obtain coordinates of one potential CNV with the maximum alternative model evidence and its score. Next, we segment the interval into 3 sections, an upstream region, a downstream region, and the segment itself. We change the baseline state for the discovered segment ($M_b:=M_a$) and continue searching for CNVs in the three regions as long as the log-likelihood ratio between alternative and base states remains large enough. We discard loci not meeting the empirically chosen thresholds and report the identified segments as a CNV if their state is different from the baseline. For more details of these methods, please refer to http://hdl.handle.net/10803/668208.

**Supplementary Results: Benchmarking**

Given the challenges associated with robust calling of CNVs genome-wide, we first set out to assess the reliability of our CNV calls. As a positive control, we compared calls at the CCL3L1 locus, a challenging multi-allelic CNV that was one of the 189 loci that passed our stringent QC, with directly measured copy number from a previous study of approximately 5000 UK Biobank participants of European ancestry (2). These 5000 participants exhibited copy numbers ranging from 0 to 5, as is typical for individuals of European ancestry (Additional file 2: Table S1). A similar distribution of copy numbers was inferred by ClinCNV in 49,953 participants from the first release of exome sequencing (Additional file 2: Table S1, Figure S1). Some of these individuals exhibited copy numbers greater than 5, but all but 15 were non-European (including 191 individuals of African ancestry, 33 of Chinese ancestry, 23 of mixed ancestry, and 8 of South Asian ancestry). In 412 individuals present in both the previous experimental study and the current ClinCNV stage 1 cohort, we found 100% concordance between copy numbers for the CCL3L1 locus (Additional file 2: Table S1).

To assess false negative rates, we ascertained common CNVs that overlapped exome capture regions in phase 3 of the 1000 genomes project. Of the 96 common CNVs identified, 60 were overlapped by a ClinCNV call of the same type and frequency (+/- 5%) (Additional file 2: Table S2).

**Figure S1**

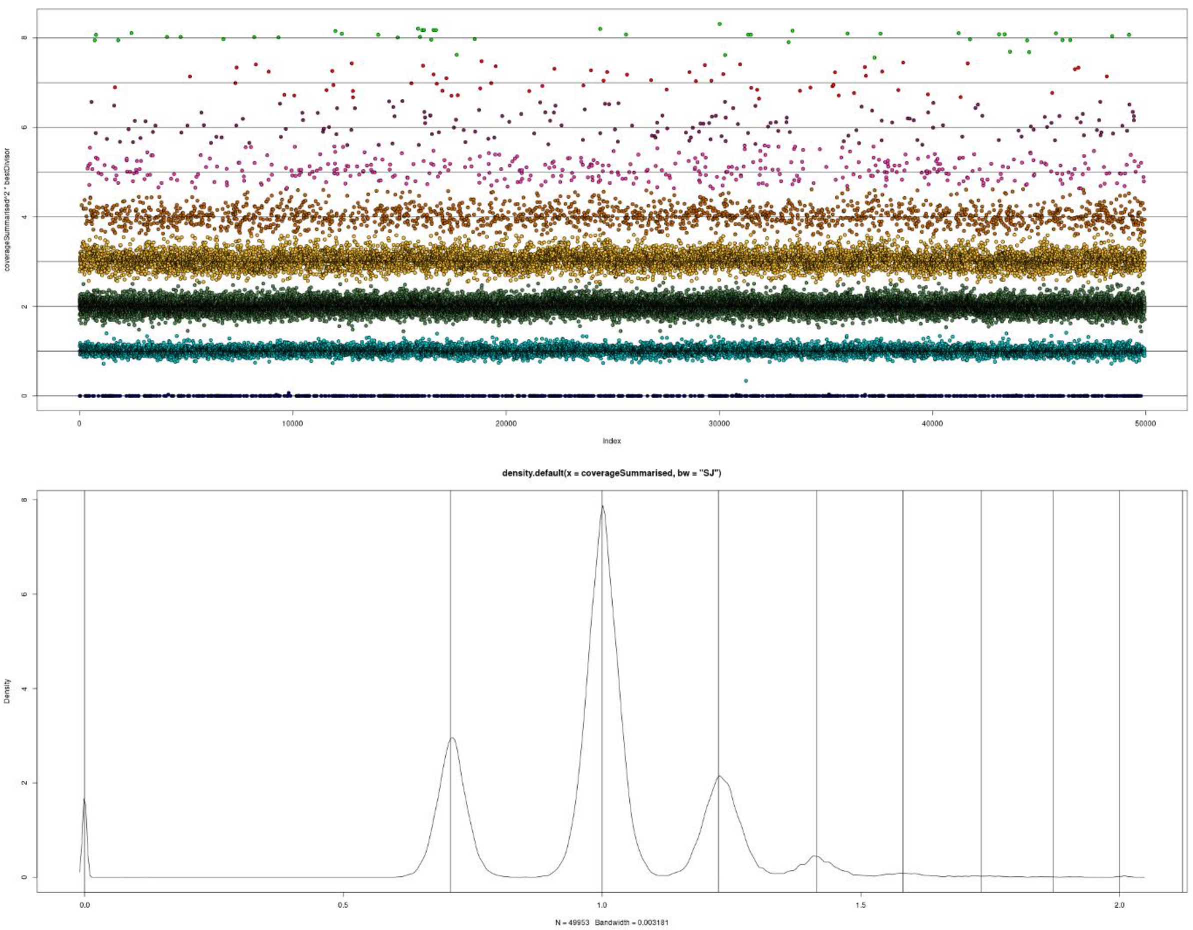


Multi-allelic CNV encompassing the CCL3L1 gene (chr17: 36195285- 36212569). The top panel shows a scatterplot of normalised coverage (y-axis) for each individual (indexed along the x-axis), and colour-coded by ClinCNV-assigned copy number. The bottom panel is a density plot of coverage values equal to sqrt( observed copy number / most frequent copy number ). So if 2 is the most frequent copy number (as is the case here), we see clustering around 0, sqrt(½), 1 and so on.

**Figure S2**

A: chr2:178432096-178444500 (stage 1 and additional stage 2 samples)

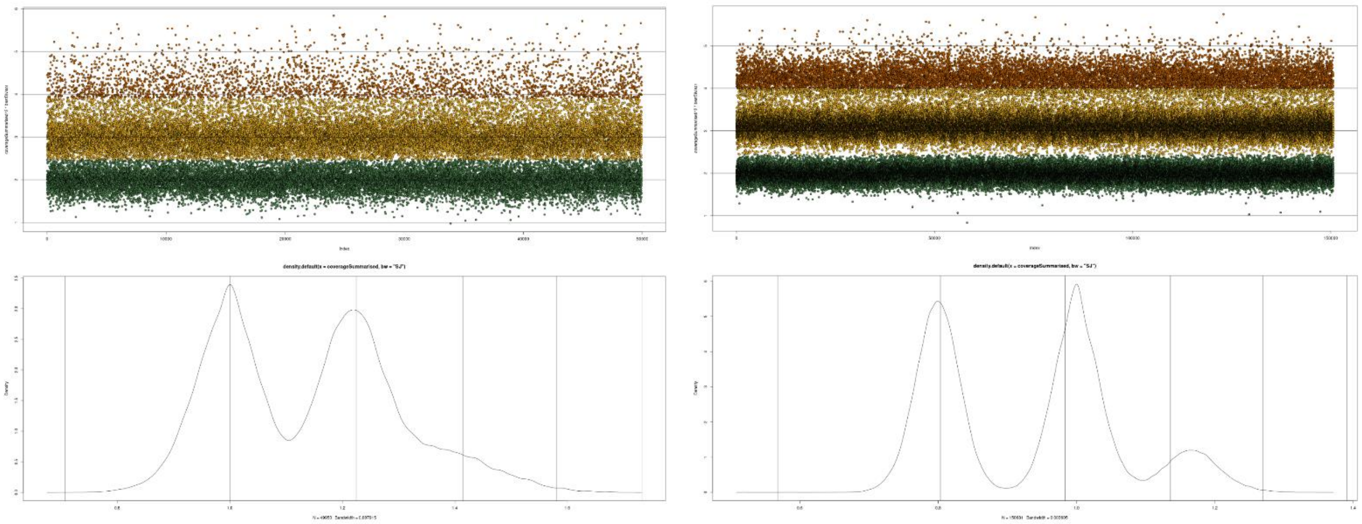

B: chr6:31026054-31027714

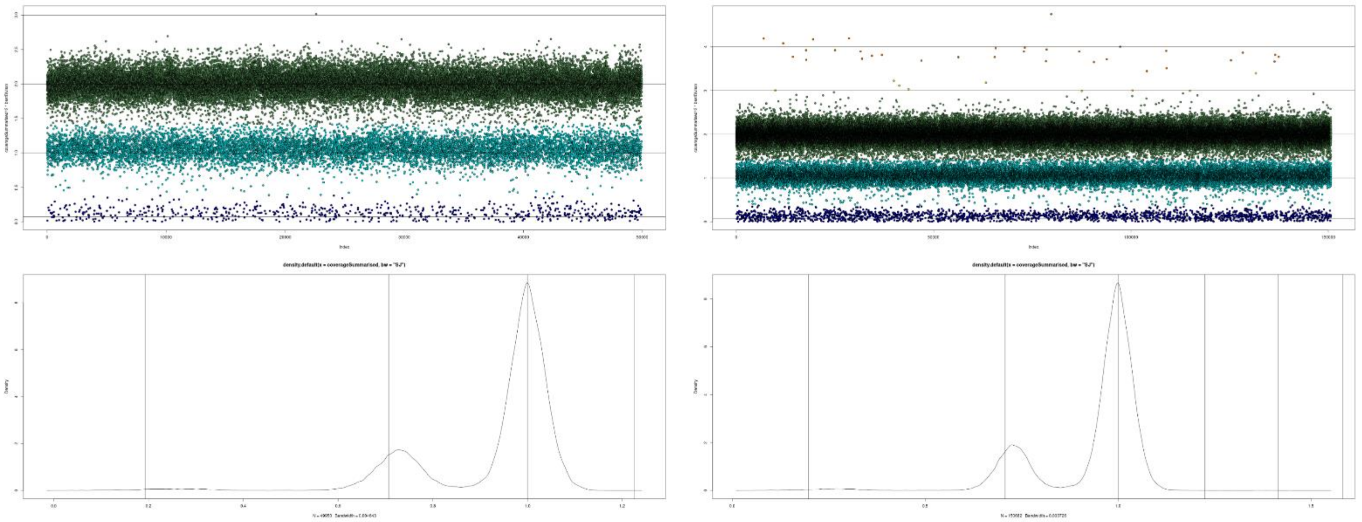

C: chr6:32641971-32666607

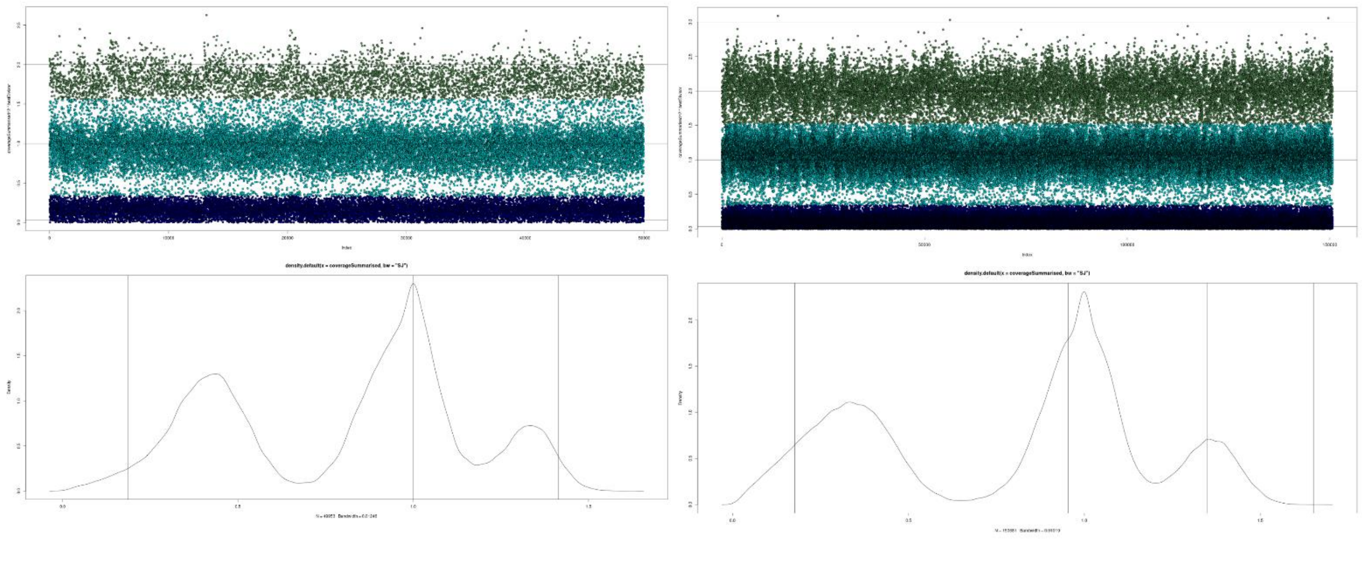

D: chr6:32827709-32828045

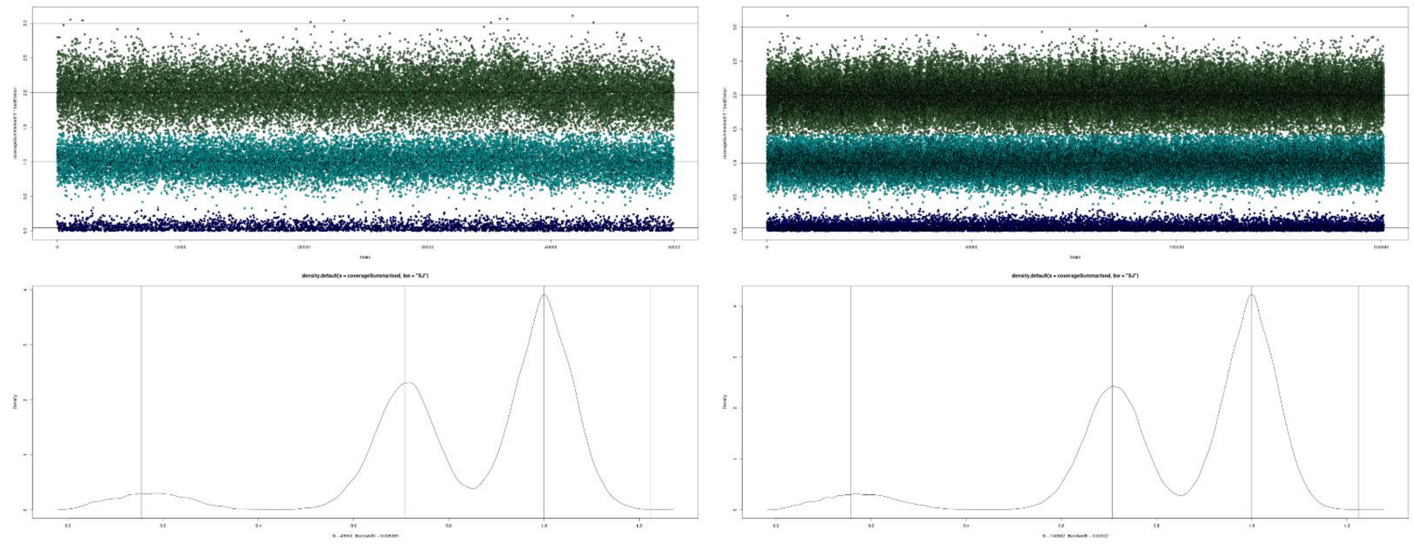


E: chr12:132507235-132511952

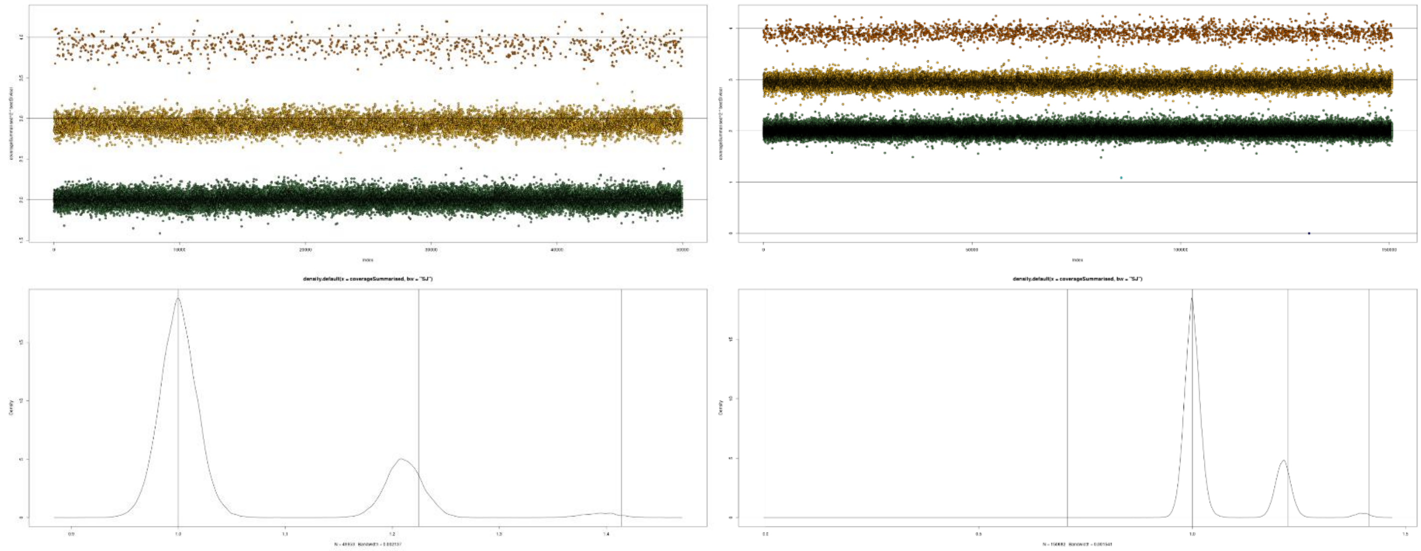

**Figure S3**

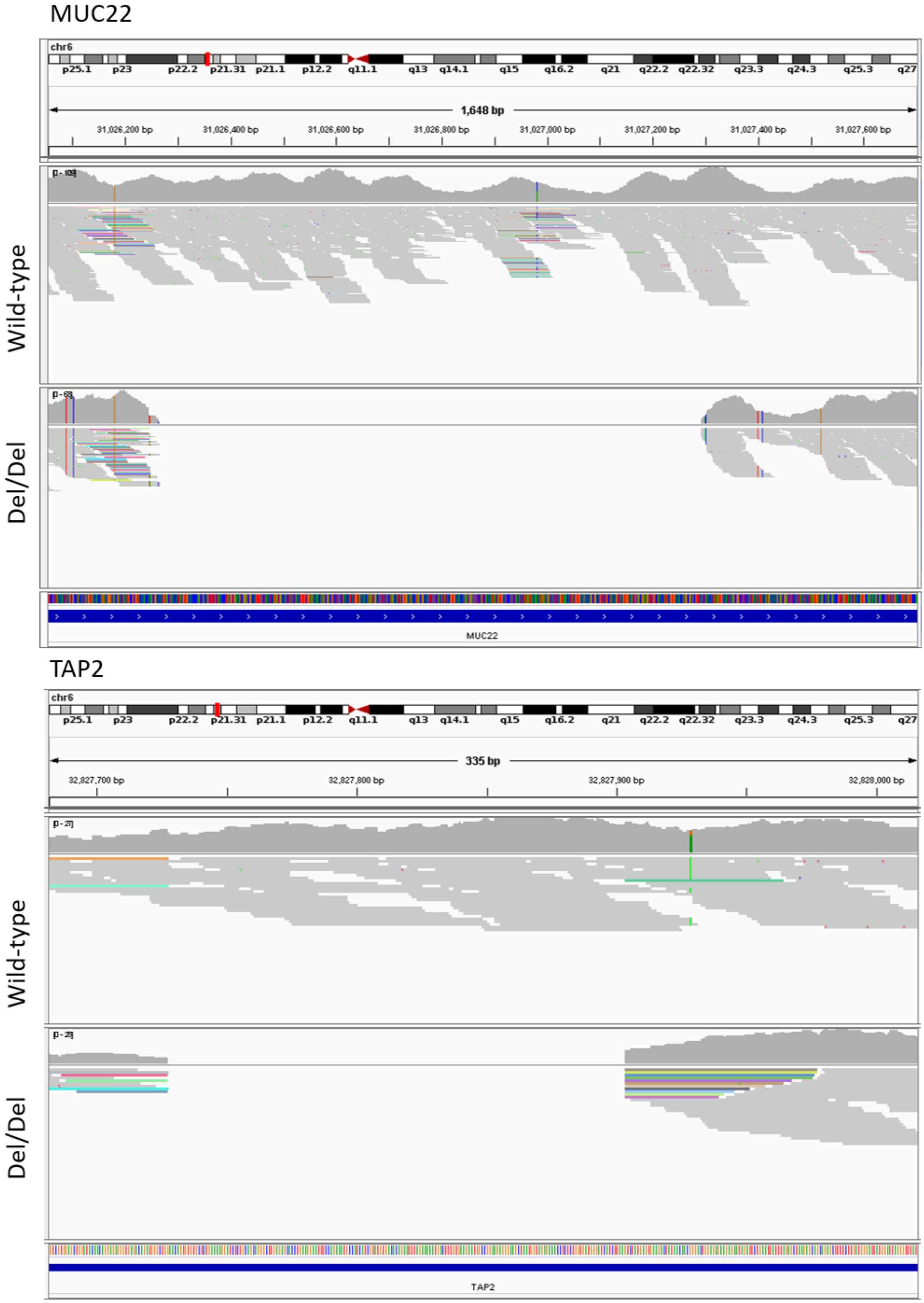
MUC22
chr6
p25.1 p23 p22.2 p21.31 p21.1 p12.2 q11.1 q13 q14.1 q15 q16.2 q21 q22.2 q22.32 q23.3 q24.3 q25.3 q27
1,648 bp
31,026,200 bp
31,026,400 bp
31,026,600 bp
31,026,800 bp
31,027,000 bp
31,027,200 bp
31,027,400 bp
31,027,600 bp
Wild-type
Del/Del
MUC22
TAP2
chr6
p25.1 p23 p22.2 p21.31 p21.1 p12.2 q11.1 q13 q14.1 q15 q16.2 q21 q22.2 q22.32 q23.3 q24.3 q25.3 q27
335 bp
32,827,700 bp
32,827,800 bp
32,827,900 bp
32,828,000 bp
Wild-type
Del/Del
TAP2

IGV images of the *MUC22* and *TAP2* CNV regions in an individual with no deletion (wild-type) and an individual homozygous for the deletion (del/del). Grey mounds in the top panels indicate read depth and grey lines in the bottom panels indicate reads from exome sequencing.

**Figure S4**

HLA-DQA1

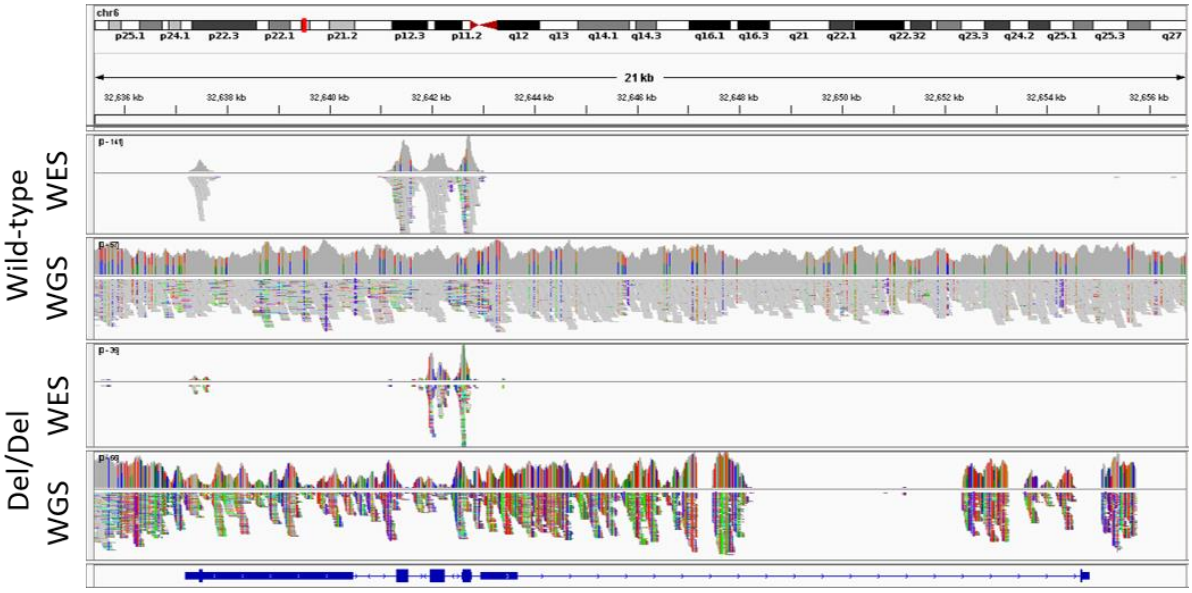


HLA-DQB1

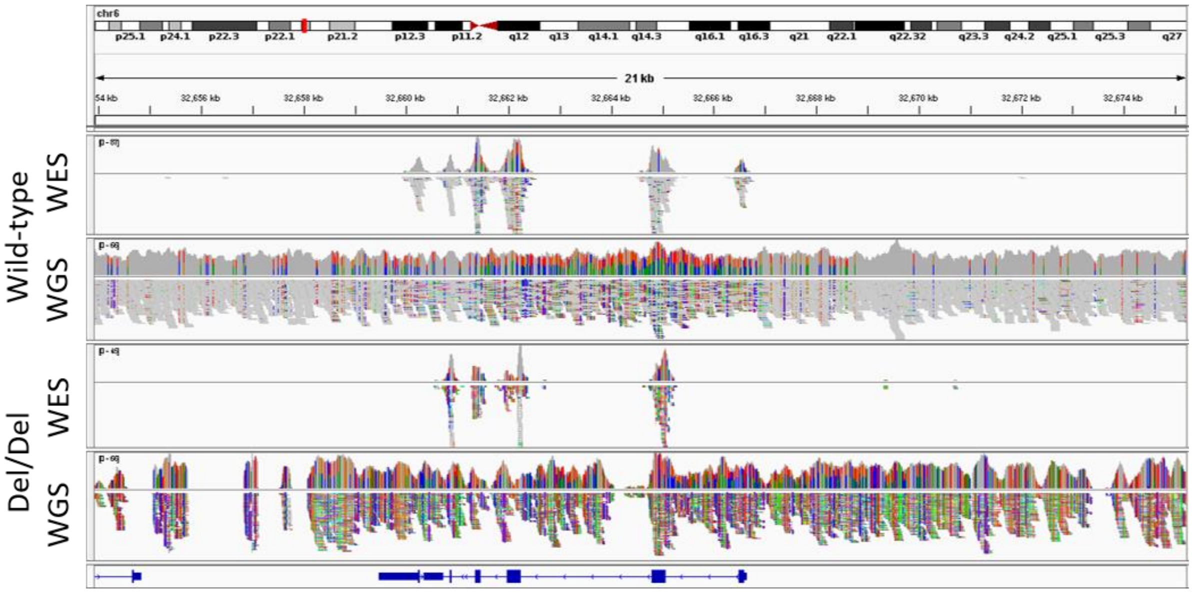


IGV images of the *HLA-DQA1/DQB1* CNV region in an individual with no deletion (wild-type) and an individual homozygous for the deletion (del/del). WES = whole-exome sequencing. WGS = pilot whole-genome sequencing. Grey mounds in the top panels indicate read depth and grey lines in the bottom panels indicate sequencing reads. Colours indicate mismatches from the reference sequence.

**Figure S5**

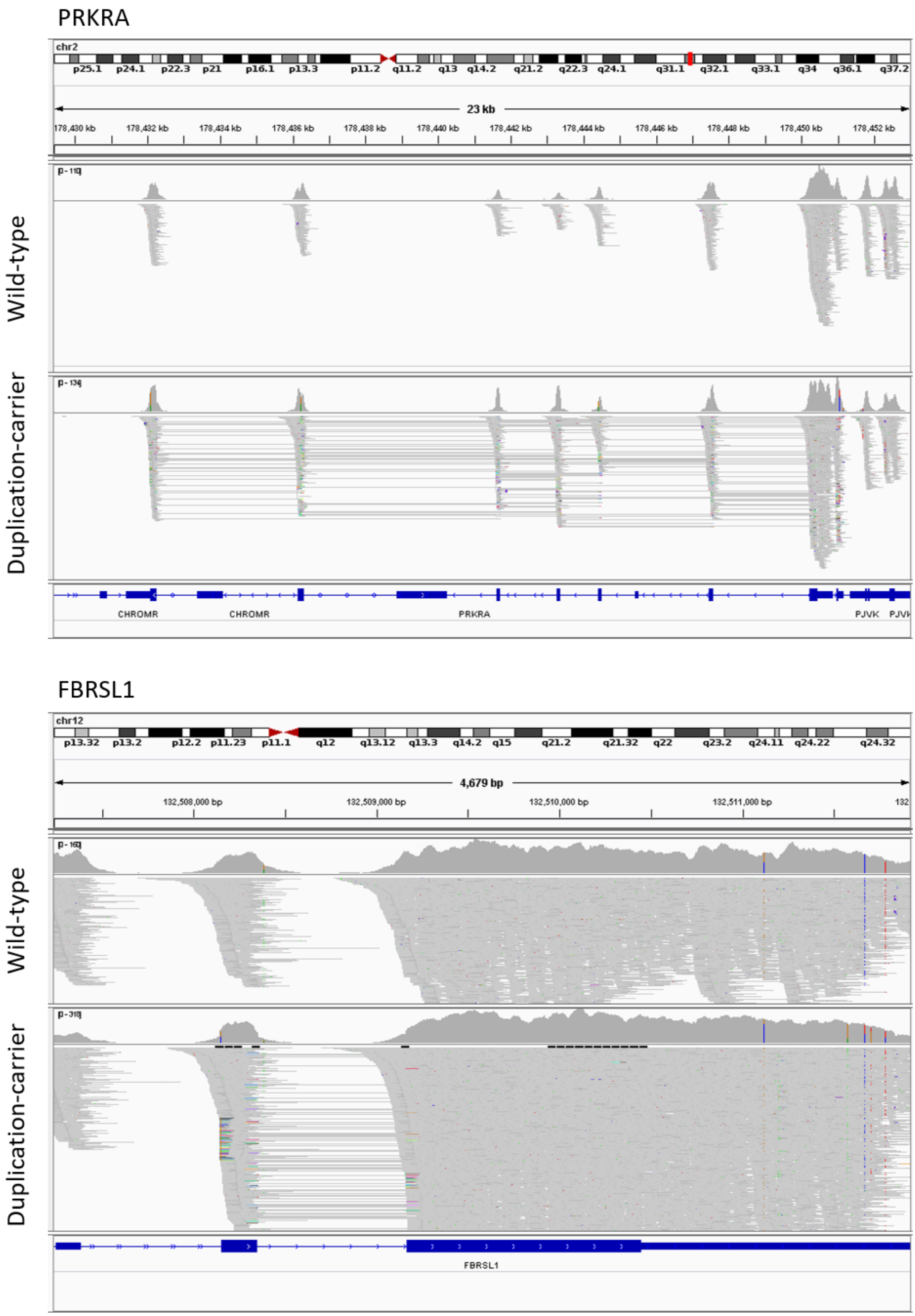
PRKRA
chr2
23 kb
178,430 kb
178,432 kb
178,434 kb
178,436 kb
178,438 kb
178,440 kb
178,442 kb
178,444 kb
178,446 kb
178,448 kb
178,450 kb
178,452 kb
Wild-type
Duplication-carrier
CHROMR
CHROMR
PRKRA
PJVK
FBRSL1
chr12
4,679 bp
132,508,000 bp
132,509,000 bp
132,510,000 bp
132,511,000 bp
Wild-type
Duplication-carrier
FBRSL1

IGV images of the *PRKRA* and *FBRSL1* CNV regions in an individual with no duplication (wild-type) and an individual carrying the duplication.  Grey mounds in the top panels indicate read depth and grey boxes joined by lines indicate read pairs from exome sequencing.
